# Views of American Democracy and Society and Support for Political Violence: First Report from a Nationwide Population-Representative Survey

**DOI:** 10.1101/2022.07.15.22277693

**Authors:** Garen J. Wintemute, Sonia Robinson, Andrew Crawford, Julia P. Schleimer, Amy Barnhorst, Vicka Chaplin, Daniel Tancredi, Elizabeth A. Tomsich, Veronica A. Pear

## Abstract

**Background:** Several social trends in the United States (US) suggest an increasing risk for political violence. Little is known about support for and personal willingness to engage in political violence and how those measures vary with lethality of violence, specific circumstances, or specific populations as targets.

**Design, Setting, Participants:** Cross-sectional nationwide survey conducted May 13 to June 2, 2022; participants were adult members of the Ipsos KnowledgePanel.

**Main Outcomes and Measures:** Weighted, population-representative proportions endorsing an array of beliefs about American democracy and society and the use of violence, including political violence, and extrapolations to the US adult population.

**Results:** The analytic sample included 8,620 respondents; 50.6% (95% Confidence Interval (CI) 49.4%, 51.7%) were female; mean (SD) age was 48.4 (18.0) years. Two-thirds of respondents (67.2%, 95% CI 66.1%, 68.4%) perceived “a serious threat to our democracy,” but more than 40% agreed that “having a strong leader for America is more important than having a democracy” and that “in America, native-born white people are being replaced by immigrants.” Half (50.1%) agreed that “in the next few years, there will be civil war in the United States.” Among 6,768 respondents who considered violence to be at least sometimes justified to achieve 1 or more specific political objectives, 12.2% were willing to commit political violence themselves “to threaten or intimidate a person,” 10.4% “to injure a person,” and 7.1% “to kill a person.” Among all respondents, 18.5% thought it at least somewhat likely that within the next few years, in a situation where they believed political violence was justified, “I will be armed with a gun”; 4.0% thought it at least somewhat likely that “I will shoot someone with a gun.”

**Conclusions and Relevance:** Coupled with prior research, these findings suggest a continuing alienation from and mistrust of American democratic society and its institutions. Substantial minorities of the population endorse violence, including lethal violence, to obtain political objectives. Efforts to prevent that violence, which a large majority of Americans already reject, should proceed rapidly based on the best evidence available. Further research will inform future prevention efforts.

## INTRODUCTION

Recent events in the United States (US)—mass shootings, Supreme Court decisions, hearings of the House committee investigating the January 6 attack on the Capitol, and others—have reminded Americans of the daily presence of violence in their nation’s public life. This study is motivated by 5 recent trends that, in their apparent convergence, create the potential for even greater violence that could put at risk the future of the US as a free and democratic society.

First is a striking rise in violence, and particularly in firearm violence. The 28% increase in homicide from 2019 to 2020^1^ was the largest single-year percentage increase ever recorded.^2^ Firearms accounted for 57.7% of violent deaths in 2019 but 62.1% in 2020, when 78.9% of homicides (19,995 of 25,356) and 52.8% of suicides (24,292 of 45,979) involved firearms.^1, 3^

Second is an equally unprecedented increase in firearm purchasing that began with the onset of the COVID-19 pandemic in January 2020 and, except for a brief respite late in 2021, has continued through June 2022. ^2, 4^ From January 2020 through June 2022, background checks on firearm purchasers have averaged 46.6% above expected levels (Supplement Figure 1); an estimated 14.5 million excess background checks have been conducted, of 45.7 million checks altogether.

Third is growing uncertainty about the stability and value of democracy in the US. Most Americans across the political spectrum now perceive a serious threat to democracy in the US.^5, 6^ At the same time, nearly 70% of adults—with very similar results for Democrats and Republicans—agree that “American democracy only serves the interests of the wealthy and powerful.”^7^ Approximately 20% of Republicans, conservatives, and voters for Donald Trump (and 9% of Democrats, liberals, and voters for Joe Biden) disagree with the statement that “democracy is [the] best form of government.”^8^

Fourth is the expansion into the mainstream of American public opinion of extreme, false beliefs about American society. Approximately 1 adult in 5 endorses the core elements of the Q-Anon belief complex, that “government, media, and financial worlds in the US are controlled by a group of Satan-worshipping pedophiles” (16%) and that “there is a storm coming soon that will sweep away the elites in power and restore the rightful leaders” (22%).^9^ Nearly 1 adult in 3 (32%) endorses the assertion that “a group of people in this country [is] trying to replace native-born Americans with immigrants.”^10^

Fifth is growing support for the use of violence to accomplish political or social objectives. More than a third (36%) of American adults (56% of Republicans and 22% of Democrats) agree that “the traditional American way of life is disappearing so fast that we may have to use force to save it.”^7^ Nearly one-fifth of adults (18%) agree that “because things have gotten so far off track, true American patriots may have to resort to violence in order to save our country.”^9^

Research on the prevalence and determinants of support for political violence in the US is sparse.^11–15^ Existing work has been criticized on multiple grounds, including failures to define violence, to determine whether support for political violence reflects support for violence generally, and to determine whether persons who endorse political violence are willing to engage in such violence themselves.^14, 15^

Many important and urgent questions remain insufficiently explored, or unexplored altogether. Does support for political violence reflect a general predisposition to violence as a means of solving problems? How prevalent are support for, and willingness to engage in, political violence when that term is defined? How do those prevalences vary with specific political objectives for which violence might be employed, with the lethality of that violence, and with its target? Beyond demographics, what individual characteristics (e.g., extreme political and social beliefs, firearm ownership) and community characteristics are associated with support for political violence? What specific preparations for political violence have its supporters made?

We developed the 2022 Life in America survey to answer these and related questions with data from a large nationally representative sample, augmented by oversamples for populations of special interest. This first report outlines the study’s overall methods and presents descriptive tabulations of data from the main study sample on key questions regarding respondents’ political and social beliefs and their support for—and willingness to engage in— political violence.

## METHODS

Data for this cross-sectional survey study are from the 2022 Life in America Survey, which was designed by the authors and administered online in English and Spanish from May 13 to June 2, 2022 by the survey research firm Ipsos.^16^ Before participants accessed the questionnaire, they were provided informed consent language that concluded, “[by] continuing, you are agreeing to participate in this study.” The study was approved by the University of California Davis Institutional Review Board and is reported following American Association for Public Opinion Research guidelines.^17^

### Participants

Respondents were drawn from the Ipsos KnowledgePanel, an online research panel that has been widely used in population-based research, including studies of violence and firearm ownership.^18–23^

To establish a nationally representative panel, members are recruited on an ongoing basis through address-based probability sampling using data from the US Postal Service’s Delivery Sequence File.^24^ Recruited adults in households without internet access are provided a web-enabled device and free internet service. A probability-proportional-to-size procedure was used to select a study-specific sample. All panel members who were aged 18 years and older were eligible for selection. Invitations were sent by e-mail; automatic reminders were delivered to non-respondents by e-mail and telephone beginning 3 days later.

A final survey weight variable provided by Ipsos adjusted for the initial probability of selection into KnowledgePanel and for survey-specific nonresponse and over- or under-coverage using design weights with post-stratification raking ratio adjustments. With weighting, the sample is designed to be statistically representative of the noninstitutionalized adult population of the US as reflected in the 2021 March supplement of the Current Population Survey.

### Measures

Sociodemographic data were collected by Ipsos as KnowledgePanel members created and maintained their member profiles. Survey questions that supplied data for this analysis covered 3 broad domains: beliefs regarding democracy and the potential for violence in the US; beliefs regarding American society and institutions; and support for and willingness to engage in violence, including political violence. Prior polls or surveys on these topics were reviewed, and selected questions were included or adapted in this questionnaire to track trends in opinion and provide context for responses to questions that had not been asked previously. The full text of all questions reported on here, including sources for questions from prior surveys, is in the Supplement.

The questionnaire used the phrase “force or violence” in place of violence. Force or violence was defined as “physical force strong enough that it could cause pain or injury to a person.” “Force or violence to advance an important political objective that you support” was used in questions about respondents’ support for and willingness to engage in political violence.

### Implementation

Ipsos translated the questionnaire into Spanish, and interpreting services staff at UC Davis Medical Center reviewed the translation. Forty KnowledgePanel members participated in a pretest of the English language version that was administered April 27 to May 2, 2022.

Respondents were randomized 1:1 to receive response options in order from negative to positive valence (e.g., from ‘do not agree’ to ‘strongly agree’) or the reverse throughout the questionnaire. Where a question presented multiple statements for respondents to consider, the order in which those statements were presented was randomized unless ordering was necessary. Logic-driving questions (those to which responses might invoke a skip pattern) included non-response prompts.

To minimize inattentive responses to questions regarding political violence, those questions were immediately preceded by a question asking respondents about the justifiability of the use of force or violence under 7 other conditions. These were presented to all respondents in a fixed order from what the authors considered likely to be seen as justifying violence (“in self-defense”) to unlikely (“to get respect”). This was done to create an expected response transition from support to nonsupport that respondents would need to reverse to indicate support for political violence. Questions on personal willingness to engage in political violence were asked only of respondents who considered violence to be at least sometimes justified to achieve 1 or more specific political objectives.

### Statistical Analysis

To generate prevalence estimates, we calculated weighted percentages and 95% confidence intervals (CI) for each measure using PROC SURVEYFREQ in SAS version 9.4 (SAS Institute, Inc., Cary, NC) and Complex Samples Frequencies in IBM SPSS Statistics, version 28 (IBM Corp., Armonk, NY). Estimated counts of adults in the US were generated by simple extrapolation, multiplying weighted percentages from our sample by the estimated adult population of the US as of July 1, 2021 (258.33 million persons).^25^

## RESULTS

Of 15,449 panel members invited to participate as part of the main study sample, 8,620 completed the survey, yielding a 55.8% completion rate. The median survey completion time was 15.7 minutes (Interquartile Range, 11.4-23.0 minutes). Item non-response ranged from 0.28% to 2.34%.

Slightly more than half of the respondents (50. 6%, 95% CI 49.4%, 51.7%) were female; 62.6% (95% CI 61.4%, 63.9%) were white, non-Hispanic; 11.9% (95% CI 11.1%, 12.8%) were Black, non-Hispanic; 16.9% (95% CI 15.9%, 17.8%) were Hispanic (of any race); and 5.4% (95% CI 4.8%, 6.1%) were Asian American/Pacific Islander, non-Hispanic (Table 1). Mean (SD) respondent age was 48.4 (18.0) years. Compared to nonrespondents, respondents were older and more frequently white, non-Hispanic; were more often married; had higher education and income; and were less likely to be working (Supplemental Table 1).

**Table 1.** Personal characteristics of respondents

| Characteristic | Respondents (n= 8,620) |  |
| --- | --- | --- |
|  | Unweighted N | Weighted %<br>(95% CI) |
| **Age** (mean [SD]) | 53.8 (17.2) | 48.4 (18.0) |
| 18-24 | 447 | 10.5 (9.6, 11.5) |
| 25-34 | 1024 | 16.6 (15.6, 17.6) |
| 35-44 | 1374 | 18.5 (17.6, 19.5) |
| 45-54 | 1215 | 14.5 (13.7, 15.3) |
| 55-64 | 1833 | 17.4 (16.6, 18.2) |
| 65-74 | 1788 | 14.4 (13.7, 15.1) |
| 75+ | 939 | 8.0 (7.4, 8.5) |
| Non-response | 0 | 0 |
| **Gender** |  |  |
| Female | 4300 | 50.6 (49.4, 51.7) |
| Male | 4159 | 47.2 (46.1, 48.4) |
| Transgender | 41 | 0.6 (0.4, 0.8) |
| Non-binary | 44 | 0.7 (0.5, 0.9) |
| Other | 20 | 0.3 (0.1, 0.4) |
| Non-response | 56 | 0.7 (0.5, 0.9) |
| **Race/Ethnicity** |  |  |
| White, non-Hispanic | 6046 | 62.6 (61.4, 63.9) |
| Black, non-Hispanic | 834 | 11.9 (11.1, 12.8) |
| Hispanic, any race | 1084 | 16.9 (15.9, 17.8) |
| American Indian or Alaska Native, non-Hispanic | 54 | 1.3 (0.9, 1.6) |
| Asian American / Pacific Islander, non-Hispanic | 313 | 5.4 (4.8, 6.1) |
| Some other race, non-Hispanic | 22 | 0.1 (0.1, 0.2) |
| 2+ Races, non-Hispanic | 267 | 1.7 (1.5, 2.0) |
| Non-response | 0 | 0 |
| **Marital status** |  |  |
| Now married | 5246 | 56.1 (54.9, 57.3) |
| Widowed | 443 | 4.0 (3.6, 4.4) |
| Divorced | 909 | 8.7 (8.1, 9.3) |
| Separated | 139 | 1.7 (1.4, 2.1) |
| Never married | 1883 | 29.5 (28.3, 30.7) |
| Non-response | 0 | 0 |
| **Education** |  |  |
| No high school diploma or GED | 542 | 9.5 (8.7, 10.4) |
| High school graduate (diploma or GED) | 2158 | 28.3 (27.2, 29.4) |
| Some college or Associate's degree | 2364 | 27.1 (26.0, 28.1) |
|  | Unweighted N | Weighted %<br>(95% CI) |
| Bachelor's degree | 1951 | 19.7 (18.8, 20.6) |
| Master's degree or higher | 1605 | 15.4 (14.7, 16.2) |
| Non-response | 0 | 0 |
| **Household Income** |  |  |
| Less than \$10,000 | 272 | 3.9 (3.4, 4.4) |
| \$10,000 to \$24,999 | 745 | 9.0 (8.3, 9.7) |
| \$25,000 to \$49,999 | 1469 | 17.0 (16.1, 17.9) |
| \$50,000 to \$74,999 | 1414 | 16.3 (15.4, 17.2) |
| \$75,000 to \$99,999 | 1214 | 13.2 (12.4, 14) |
| \$100,000 to \$149,999 | 1500 | 17.9 (16.9, 18.8) |
| \$150,000 or more | 2006 | 22.8 (21.8, 23.7) |
| Non-response | 0 | 0 |
| **Employment** |  |  |
| Working – as a paid employee | 4323 | 54.3 (53.1, 55.4) |
| Working – self-employed | 694 | 8.0 (7.3, 8.6) |
| Not working – on temporary layoff from a job | 40 | 0.6 (0.4, 0.8) |
| Not working – looking for work | 312 | 5.1 (4.5, 5.7) |
| Not working – retired | 2478 | 20.9 (20.1, 21.8) |
| Not working – disabled | 314 | 4.2 (3.7, 4.7) |
| Not working – other | 459 | 7.0 (6.3, 7.7) |
| Non-response | 0 | 0 |
| **Census region** |  |  |
| New England | 412 | 4.7 (4.2, 5.2) |
| Mid-Atlantic | 1090 | 12.5 (11.8, 13.3) |
| East-North Central | 1267 | 14.3 (13.5, 15.1) |
| West-North Central | 604 | 6.4 (5.8, 6.9) |
| South Atlantic | 1714 | 20.5 (19.5, 21.4) |
| East-South Central | 465 | 5.8 (5.3, 6.4) |
| West-South Central | 904 | 12.0 (11.1, 12.8) |
| Mountain | 745 | 7.7 (7.1, 8.2) |
| Pacific | 1419 | 16.2 (15.3, 17.1) |
| Non-response | 0 | 0 |

### Democracy and the Potential for Violence

More than two-thirds of respondents (67.2%, 95% CI 66.1%-68.4%) perceived “a serious threat to our democracy,” and 88.8% believed it is very or extremely important “for the United States to remain a democracy” (Table 2). But at the same time, 42.4% agreed with the statement that “having a strong leader for America is more important than having a democracy”; 19.0% agreed strongly or very strongly.

Significant minorities of respondents agreed strongly or very strongly with each of 3 statements about potential conditions in the US might justify violence (Table 2): to “protect American democracy” if “elected leaders will not” (18.7%); to save “our American way of life,” which is “disappearing” (16.1%); and to “save our country” because “things have gotten so far off track” (8.1%). Half the respondents (50.1%) agreed at least somewhat that “in the next few years, there will be civil war in the United States.”

**Table 2.** Views on democracy and the potential for violence in the United States

| Statement | Respondents (n= 8,620) |  | Estimated N of adults in US |
| --- | --- | --- | --- |
|  | Unweighted N | Weighted %<br>(95% CI) | N (95% CI) (in millions) |
| When thinking about democracy in the United States these days, do you believe... |  |  |  |
| There is a serious threat to our democracy. | 6117 | 67.2 (66.1, 68.4) | 173.7 (170.7, 176.7) |
| There may be a threat to our democracy, but it is not serious. | 1832 | 23.6 (22.5, 24.6) | 60.9 (58.2, 63.7) |
| There is no threat to our democracy. | 573 | 7.8 (7.1, 8.5) | 20.1 (18.3, 21.9) |
| Non-response | 98 | 1.4 (1.1, 1.7) | 3.6 (2.8, 4.4) |
| How important do you think it is for the United States to remain a democracy? |  |  |  |
| Not important | 145 | 2.2 (1.8, 2.6) | 5.6 (4.6, 6.6) |
| Somewhat important | 510 | 7.8 (7.1, 8.5) | 20.1 (18.3, 22) |
| Very important | 1828 | 24.1 (23.1, 25.2) | 62.4 (59.6, 65.1) |
| Extremely important | 6058 | 64.7 (63.6, 65.9) | 167.2 (164.2, 170.3) |
| Non-response | 79 | 1.1 (0.9, 1.4) | 2.9 (2.2, 3.7) |
| Having a strong leader for America is more important than having a democracy. |  |  |  |
| Do not agree | 5141 | 56.0 (54.8, 57.2) | 144.7 (141.6, 147.8) |
| Somewhat agree | 1835 | 23.4 (22.3, 24.4) | 60.4 (57.7, 63.1) |
| Strongly agree | 821 | 10.3 (9.5, 11.0) | 26.5 (24.6, 28.5) |
| Very strongly agree | 702 | 8.7 (8.0, 9.4) | 22.4 (20.6, 24.2) |
| Non-response | 121 | 1.7 (1.3, 2.0) | 4.3 (3.5, 5.2) |
| If elected leaders will not protect American democracy, the people must do it themselves, even if it requires taking violent actions. |  |  |  |
| Do not agree | 4504 | 50.0 (48.9, 51.2) | 129.3 (126.2, 132.4) |
| Somewhat agree | 2468 | 29.6 (28.5, 30.7) | 76.4 (73.6, 79.2) |
|  | Unweighted N | Weighted %<br>(95% CI) | N (95% CI) (in millions) |
| Strongly agree | 834 | 10.3 (9.6, 11.1) | 26.6 (24.7, 28.6) |
| Very strongly agree | 687 | 8.4 (7.7, 9.1) | 21.7 (19.9, 23.5) |
| Non-response | 127 | 1.7 (1.4, 2) | 4.3 (3.5, 5.1) |
| Our American way of life is disappearing so fast that we may have to use force to save it. |  |  |  |
| Do not agree | 4959 | 55.6 (54.4, 56.8) | 143.7 (140.6, 146.8) |
| Somewhat agree | 2222 | 26.7 (25.7, 27.8) | 69.1 (66.3, 71.8) |
| Strongly agree | 730 | 8.9 (8.2, 9.6) | 23.0 (21.2, 24.9) |
| Very strongly agree | 585 | 7.2 (6.5, 7.8) | 18.5 (16.9, 20.2) |
| Non-response | 124 | 1.5 (1.2, 1.8) | 4.0 (3.2, 4.8) |
| Because things have gotten so far off track, true American patriots may have to resort to violence in order to save our country. |  |  |  |
| Do not agree | 6404 | 72.4 (71.3, 73.5) | 187 (184.2, 189.9) |
| Somewhat agree | 1423 | 17.6 (16.6, 18.5) | 45.4 (43, 47.8) |
| Strongly agree | 369 | 4.4 (3.9, 4.9) | 11.4 (10.1, 12.6) |
| Very strongly agree | 279 | 3.7 (3.2, 4.2) | 9.6 (8.3, 10.9) |
| Non-response | 145 | 1.9 (1.6, 2.2) | 4.9 (4, 5.8) |
| In the next few years, there will be civil war in the United States. |  |  |  |
| Do not agree | 4268 | 47.8 (46.6, 48.9) | 123.4 (120.3, 126.4) |
| Somewhat agree | 3126 | 36.4 (35.3, 37.6) | 94.1 (91.1, 97.0) |
| Strongly agree | 654 | 8.4 (7.7, 9.1) | 21.8 (20, 23.6) |
| Very strongly agree | 411 | 5.3 (4.8, 5.9) | 13.7 (12.3, 15.2) |
| Non-response | 161 | 2.1 (1.7, 2.4) | 5.4 (4.5, 6.3) |

### American Society and Institutions

Four items explored beliefs on race and “great replacement” thinking (Table 3). While 39.0% of respondents agreed strongly or very strongly that “white people benefit from advantages in society that Black people do not have,” 27.1% agreed strongly or very strongly that “discrimination against whites is as big a problem as discrimination against Blacks and other minorities.” Nearly 1 in 5 (18.6%) disagreed with the statement that “having more Black Americans, Latinos, and Asian Americans is good for the country,” and 41.2% agreed—16.2% agreed strongly or very strongly—with the proposition that “in America, native-born white people are being replaced by immigrants.”

**Table 3.**
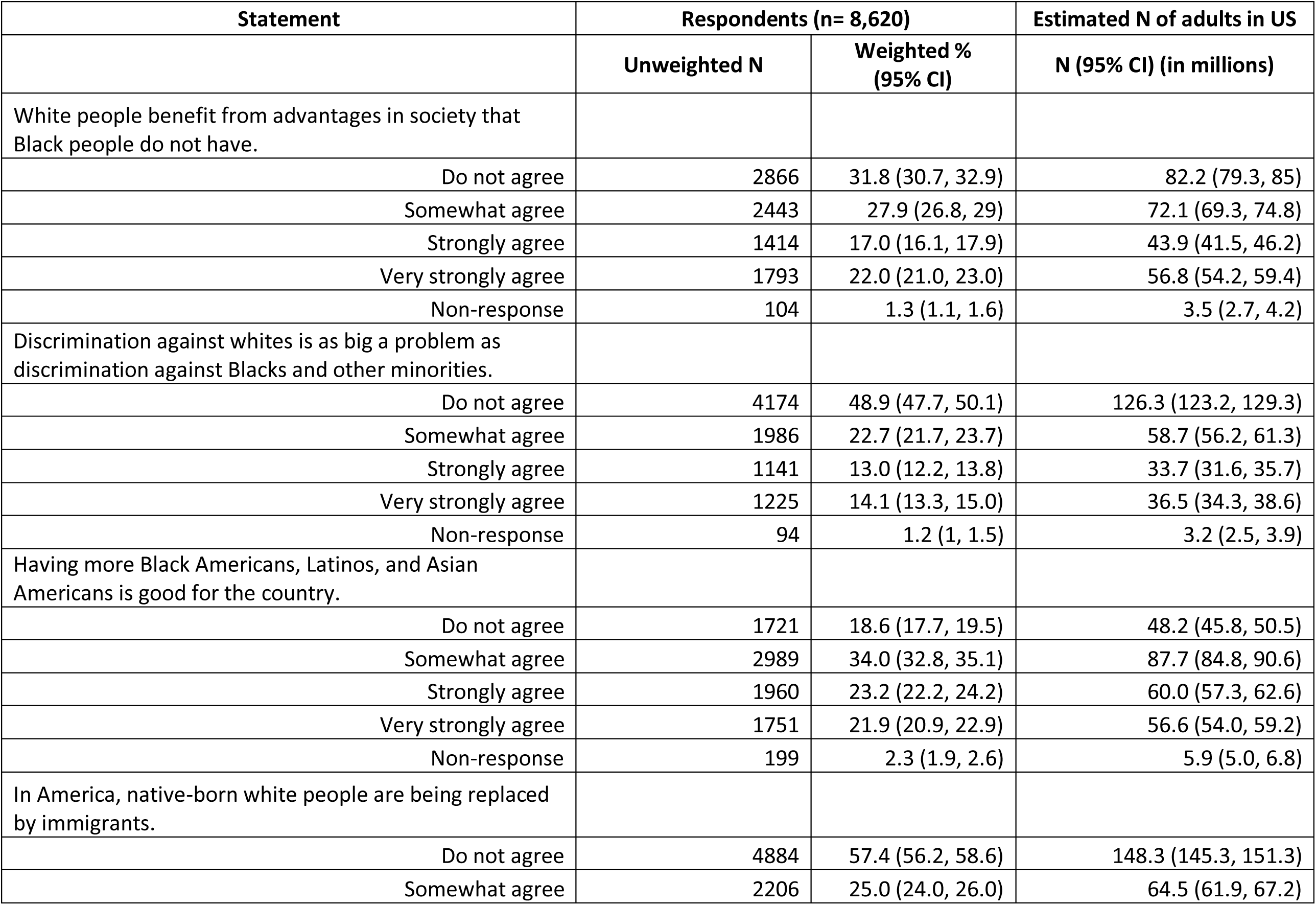
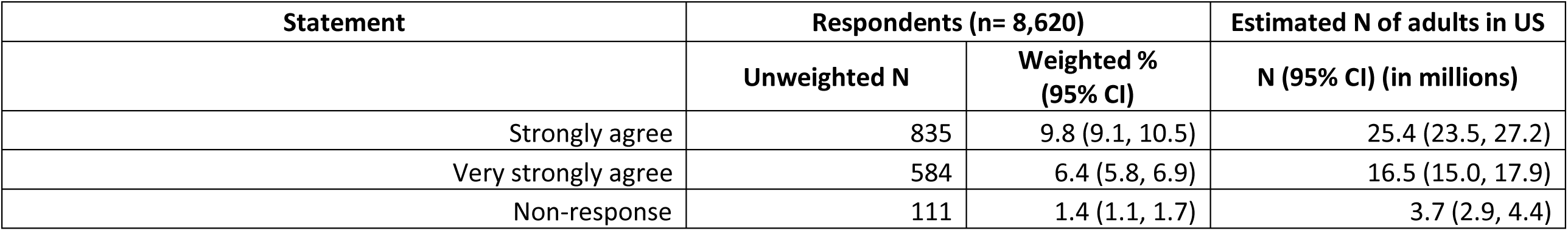
Views on race and “replacement”

| Statement | Respondents (n= 8,620) |  | Estimated N of adults in US |
| --- | --- | --- | --- |
|  | Unweighted N | Weighted %<br>(95% CI) | N (95% CI) (in millions) |
| White people benefit from advantages in society that Black people do not have. |  |  |  |
| Do not agree | 2866 | 31.8 (30.7, 32.9) | 82.2 (79.3, 85) |
| Somewhat agree | 2443 | 27.9 (26.8, 29) | 72.1 (69.3, 74.8) |
| Strongly agree | 1414 | 17.0 (16.1, 17.9) | 43.9 (41.5, 46.2) |
| Very strongly agree | 1793 | 22.0 (21.0, 23.0) | 56.8 (54.2, 59.4) |
| Non-response | 104 | 1.3 (1.1, 1.6) | 3.5 (2.7, 4.2) |
| Discrimination against whites is as big a problem as discrimination against Blacks and other minorities. |  |  |  |
| Do not agree | 4174 | 48.9 (47.7, 50.1) | 126.3 (123.2, 129.3) |
| Somewhat agree | 1986 | 22.7 (21.7, 23.7) | 58.7 (56.2, 61.3) |
| Strongly agree | 1141 | 13.0 (12.2, 13.8) | 33.7 (31.6, 35.7) |
| Very strongly agree | 1225 | 14.1 (13.3, 15.0) | 36.5 (34.3, 38.6) |
| Non-response | 94 | 1.2 (1, 1.5) | 3.2 (2.5, 3.9) |
| Having more Black Americans, Latinos, and Asian Americans is good for the country. |  |  |  |
| Do not agree | 1721 | 18.6 (17.7, 19.5) | 48.2 (45.8, 50.5) |
| Somewhat agree | 2989 | 34.0 (32.8, 35.1) | 87.7 (84.8, 90.6) |
| Strongly agree | 1960 | 23.2 (22.2, 24.2) | 60.0 (57.3, 62.6) |
| Very strongly agree | 1751 | 21.9 (20.9, 22.9) | 56.6 (54.0, 59.2) |
| Non-response | 199 | 2.3 (1.9, 2.6) | 5.9 (5.0, 6.8) |
| In America, native-born white people are being replaced by immigrants. |  |  |  |
| Do not agree | 4884 | 57.4 (56.2, 58.6) | 148.3 (145.3, 151.3) |
| Somewhat agree | 2206 | 25.0 (24.0, 26.0) | 64.5 (61.9, 67.2) |
|  | Unweighted N | Weighted %<br>(95% CI) | N (95% CI) (in millions) |
| Strongly agree | 835 | 9.8 (9.1, 10.5) | 25.4 (23.5, 27.2) |
| Very strongly agree | 584 | 6.4 (5.8, 6.9) | 16.5 (15.0, 17.9) |
| Non-response | 111 | 1.4 (1.1, 1.7) | 3.7 (2.9, 4.4) |

Another 4 items addressed the central elements of QAnon mythology and other beliefs (Table 4). More than 1 in 5 respondents (22.7%) endorsed the statement, with 9% agreeing strongly or very strongly, that US institutions are “controlled by a group of Satan-worshipping pedophiles” who traffic children for sex. Nearly 30% (29.7%) agreed (10.1% strongly or very strongly) that “a storm coming soon” will “sweep away the elites in power and restore the rightful leaders.” More than 2 in 5 (43.4%) agreed (19.3% strongly or very strongly) that “we are living in what the Bible calls ‘the end times.’” Nearly a third of respondents (32.1%) endorsed the statement that “the 2020 election was stolen from Donald Trump, and Joe Biden is an illegitimate president”; nearly 1 in 5 (18.4%) agreed strongly or very strongly.

**Table 4.** Views on American society and institutions

| Statement | Respondents (n= 8,620) |  | Estimated N of adults in US |
| --- | --- | --- | --- |
|  | Unweighted N | Weighted %<br>(95% CI) | N (95% CI) (in millions) |
| The government, media, and financial worlds in the U.S. are controlled by a group of Satan-worshipping pedophiles who run a global child sex trafficking operation. |  |  |  |
| Do not agree | 6775 | 74.9 (73.8, 76) | 193.4 (190.6, 196.2) |
| Somewhat agree | 1000 | 13.7 (12.8, 14.6) | 35.3 (33.0, 37.6) |
| Strongly agree | 329 | 4.5 (4.0, 5.1) | 11.7 (10.3, 13.1) |
| Very strongly agree | 328 | 4.5 (4.0, 5.1) | 11.7 (10.3, 13.1) |
| Non-response | 188 | 2.4 (2, 2.8) | 6.2 (5.2, 7.2) |
| There is a storm coming soon that will sweep away the elites in power and restore the rightful leaders. |  |  |  |
| Do not agree | 6031 | 67.8 (66.7, 68.9) | 175.1 (172.2, 178.1) |
| Somewhat agree | 1610 | 19.6 (18.6, 20.6) | 50.6 (48.1, 53.1) |
| Strongly agree | 429 | 5.5 (4.9, 6) | 14.1 (12.6, 15.6) |
| Very strongly agree | 348 | 4.6 (4, 5.1) | 11.8 (10.4, 13.2) |
| Non-response | 202 | 2.6 (2.2, 3) | 6.7 (5.6, 7.7) |
| The chaos in America today is evidence that we are living in what the Bible calls “the end times.” |  |  |  |
| Do not agree | 4905 | 54.7 (53.5, 55.9) | 141.4 (138.3, 144.5) |
| Somewhat agree | 2056 | 24.1 (23.1, 25.2) | 62.4 (59.7, 65.0) |
| Strongly agree | 694 | 8.9 (8.2, 9.6) | 23 (21.1, 24.8) |
| Very strongly agree | 821 | 10.4 (9.6, 11.2) | 26.9 (24.9, 28.8) |
| Non-response | 144 | 1.8 (1.5, 2.2) | 4.7 (3.9, 5.6) |
| The 2020 election was stolen from Donald Trump, and Joe Biden is an illegitimate president. |  |  |  |
|  | Unweighted N | Weighted %<br>(95% CI) | N (95% CI) (in millions) |
| Do not agree | 5761 | 66.2 (65.1, 67.4) | 171.1 (168.2, 174.0) |
| Somewhat agree | 1142 | 13.7 (12.9, 14.5) | 35.4 (33.2, 37.6) |
| Strongly agree | 498 | 5.9 (5.4, 6.5) | 15.3 (13.9, 16.8) |
| Very strongly agree | 1083 | 12.5 (11.7, 13.2) | 32.2 (30.2, 34.2) |
| Non-response | 136 | 1.7 (1.4, 2.0) | 4.3 (3.5, 5.1) |

### Violence

Respondents’ views on the justifiability of violence varied substantially, and predictably, with circumstance (Figure 1). Nearly all respondents saw violence as at least sometimes justified in self-defense (97.1%), or to prevent assaultive (96.5%) or self-inflicted (92.8%) injury to others. Conversely, large majorities reported that violence to win an argument (85.7%), respond to an insult (81.5%), or get respect (86.2%) was never justified. One in 5 respondents (20.5%) believed that “in general,” political violence was at least sometimes justified; 3.0% considered it usually or always justified (Table 5, Figure 1).

**Figure 1.**
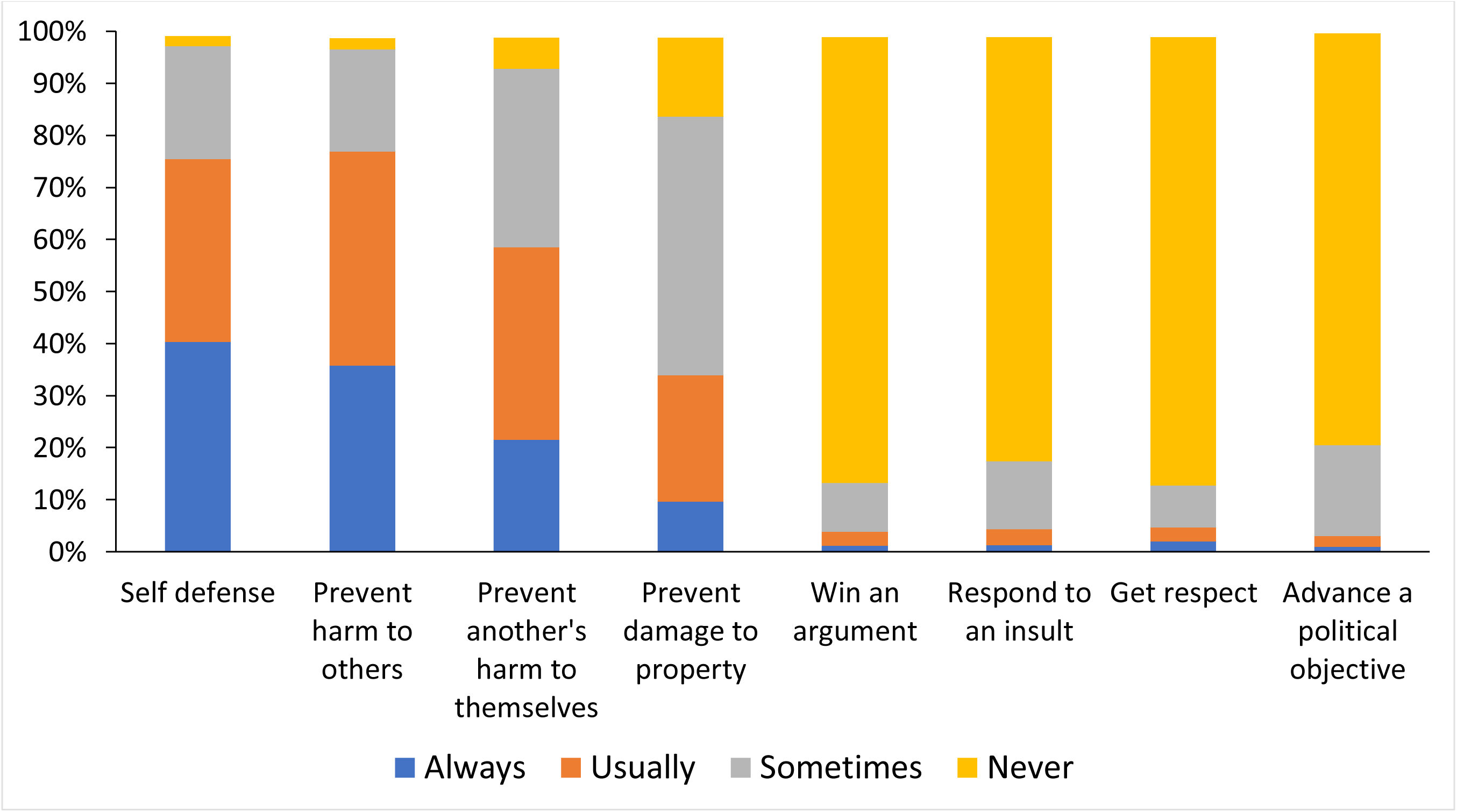
Justifiability of use or force or violence in specific situations Respondents (n= 8,620) were asked, “What do you think about the use of force or violence in the following situations?” with response options always/usually/sometimes/never justified.

**Table 5.**
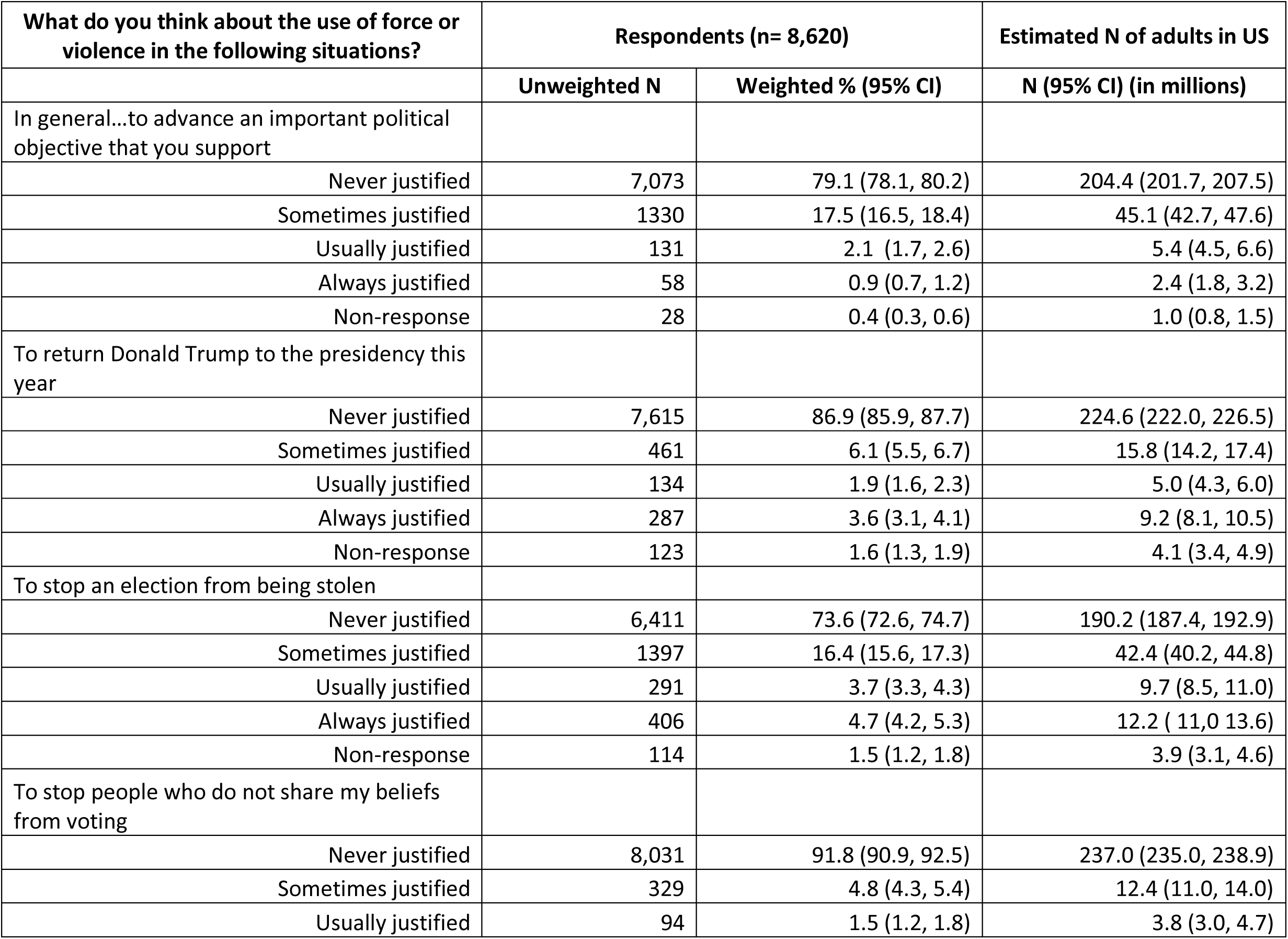
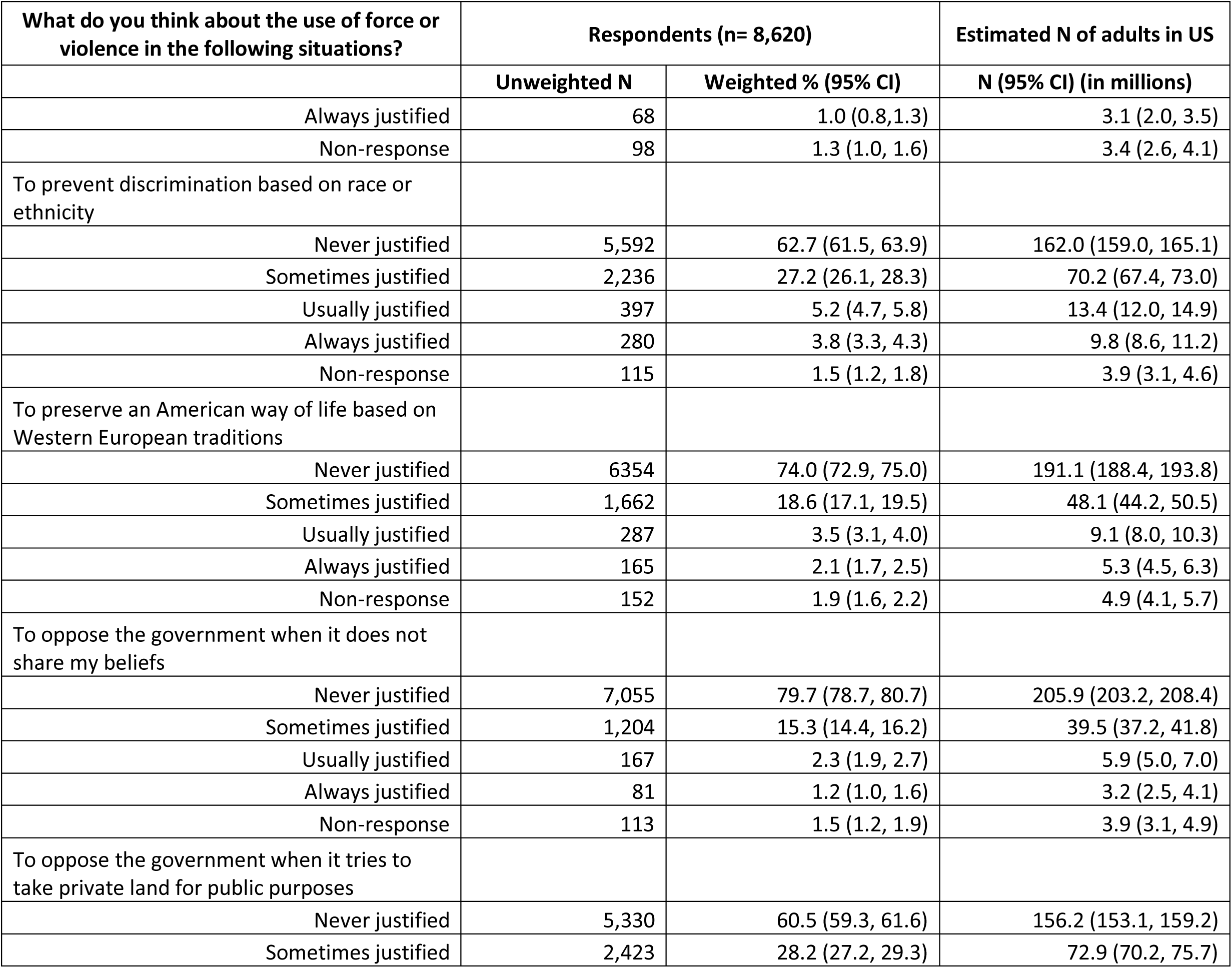
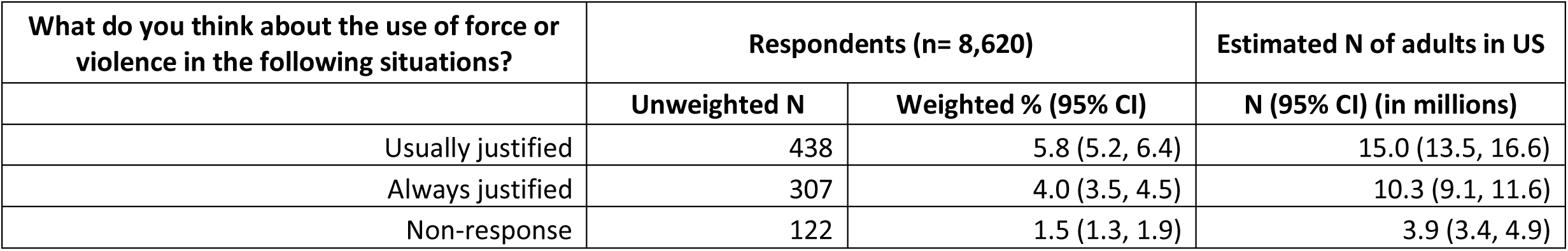
Views on political violence, generally and by circumstance

| What do you think about the use of force or violence in the following situations? | Respondents (n= 8,620) |  | Estimated N of adults in US |
| --- | --- | --- | --- |
|  | Unweighted N | Weighted % (95% CI) | N (95% CI) (in millions) |
| In general...to advance an important political objective that you support |  |  |  |
| Never justified | 7,073 | 79.1 (78.1, 80.2) | 204.4 (201.7, 207.5) |
| Sometimes justified | 1330 | 17.5 (16.5, 18.4) | 45.1 (42.7, 47.6) |
| Usually justified | 131 | 2.1 (1.7, 2.6) | 5.4 (4.5, 6.6) |
| Always justified | 58 | 0.9 (0.7, 1.2) | 2.4 (1.8, 3.2) |
| Non-response | 28 | 0.4 (0.3, 0.6) | 1.0 (0.8, 1.5) |
| To return Donald Trump to the presidency this year |  |  |  |
| Never justified | 7,615 | 86.9 (85.9, 87.7) | 224.6 (222.0, 226.5) |
| Sometimes justified | 461 | 6.1 (5.5, 6.7) | 15.8 (14.2, 17.4) |
| Usually justified | 134 | 1.9 (1.6, 2.3) | 5.0 (4.3, 6.0) |
| Always justified | 287 | 3.6 (3.1, 4.1) | 9.2 (8.1, 10.5) |
| Non-response | 123 | 1.6 (1.3, 1.9) | 4.1 (3.4, 4.9) |
| To stop an election from being stolen |  |  |  |
| Never justified | 6,411 | 73.6 (72.6, 74.7) | 190.2 (187.4, 192.9) |
| Sometimes justified | 1397 | 16.4 (15.6, 17.3) | 42.4 (40.2, 44.8) |
| Usually justified | 291 | 3.7 (3.3, 4.3) | 9.7 (8.5, 11.0) |
| Always justified | 406 | 4.7 (4.2, 5.3) | 12.2 (11.0, 13.6) |
| Non-response | 114 | 1.5 (1.2, 1.8) | 3.9 (3.1, 4.6) |
| To stop people who do not share my beliefs from voting |  |  |  |
| Never justified | 8,031 | 91.8 (90.9, 92.5) | 237.0 (235.0, 238.9) |
| Sometimes justified | 329 | 4.8 (4.3, 5.4) | 12.4 (11.0, 14.0) |
| Usually justified | 94 | 1.5 (1.2, 1.8) | 3.8 (3.0, 4.7) |
|  | Unweighted N | Weighted % (95% CI) | N (95% CI) (in millions) |
| Always justified | 68 | 1.0 (0.8, 1.3) | 3.1 (2.0, 3.5) |
| Non-response | 98 | 1.3 (1.0, 1.6) | 3.4 (2.6, 4.1) |
| To prevent discrimination based on race or ethnicity |  |  |  |
| Never justified | 5,592 | 62.7 (61.5, 63.9) | 162.0 (159.0, 165.1) |
| Sometimes justified | 2,236 | 27.2 (26.1, 28.3) | 70.2 (67.4, 73.0) |
| Usually justified | 397 | 5.2 (4.7, 5.8) | 13.4 (12.0, 14.9) |
| Always justified | 280 | 3.8 (3.3, 4.3) | 9.8 (8.6, 11.2) |
| Non-response | 115 | 1.5 (1.2, 1.8) | 3.9 (3.1, 4.6) |
| To preserve an American way of life based on Western European traditions |  |  |  |
| Never justified | 6354 | 74.0 (72.9, 75.0) | 191.1 (188.4, 193.8) |
| Sometimes justified | 1,662 | 18.6 (17.1, 19.5) | 48.1 (44.2, 50.5) |
| Usually justified | 287 | 3.5 (3.1, 4.0) | 9.1 (8.0, 10.3) |
| Always justified | 165 | 2.1 (1.7, 2.5) | 5.3 (4.5, 6.3) |
| Non-response | 152 | 1.9 (1.6, 2.2) | 4.9 (4.1, 5.7) |
| To oppose the government when it does not share my beliefs |  |  |  |
| Never justified | 7,055 | 79.7 (78.7, 80.7) | 205.9 (203.2, 208.4) |
| Sometimes justified | 1,204 | 15.3 (14.4, 16.2) | 39.5 (37.2, 41.8) |
| Usually justified | 167 | 2.3 (1.9, 2.7) | 5.9 (5.0, 7.0) |
| Always justified | 81 | 1.2 (1.0, 1.6) | 3.2 (2.5, 4.1) |
| Non-response | 113 | 1.5 (1.2, 1.9) | 3.9 (3.1, 4.9) |
| To oppose the government when it tries to take private land for public purposes |  |  |  |
| Never justified | 5,330 | 60.5 (59.3, 61.6) | 156.2 (153.1, 159.2) |
| Sometimes justified | 2,423 | 28.2 (27.2, 29.3) | 72.9 (70.2, 75.7) |
|  | Unweighted N | Weighted % (95% CI) | N (95% CI) (in millions) |
| Usually justified | 438 | 5.8 (5.2, 6.4) | 15.0 (13.5, 16.6) |
| Always justified | 307 | 4.0 (3.5, 4.5) | 10.3 (9.1, 11.6) |
| Non-response | 122 | 1.5 (1.3, 1.9) | 3.9 (3.4, 4.9) |

Substantial minorities of respondents considered violence to be at least sometimes justified to achieve a wide array of specific political objectives (Table 5, Supplemental Figure 2): 11.6% “to return Donald Trump to the presidency this year,” 24.8% “to stop an election from being stolen,” 7.3% “to stop people who do not share my beliefs from voting,” 24.2% “to preserve an American way of life based on Western European traditions,” 18.8% “to oppose the government when it does not share my beliefs,” and 38.0% “to oppose the government when it tries to take private land for public purposes.” More than a third of respondents (36.2%) reported that violence was at least sometimes justified “to prevent discrimination based on race or ethnicity.”

The 6,768 respondents who considered violence to be at least sometimes justified to achieve 1 or more specific political objectives were presented 3 series of items regarding personal willingness to use force or violence “in a situation where you think force or violence is justified to advance an important political objective.” One series (Table 6) concerned types of violence. Among these respondents, 13.7% were at least somewhat willing to use force or violence “to damage property,” 12.2% “to threaten or intimidate a person,” 10.4% “to injure a person,” and 7.1% “to kill a person.” Approximately 3% were very or completely willing to threaten, injure, or kill another person to advance a political objective.

**Table 6.** Personal willingness to engage in specific forms of political violence among respondents who considered violence to be at least sometimes justified to achieve 1 or more specific political objectives

| In a situation where you think force or violence is justified to advance an important political objective... | Unweighted N | Weighted % (95% CI) |  | Estimated N of adults in US |
| --- | --- | --- | --- | --- |
|  |  | Among respondents asked the question (n= 6,768)* | Among the full sample (n= 8,620)** | N (95% CI) (in millions) |
| How willing would <u>you personally</u> be to use force or violence in each of these ways? |  |  |  |  |
| To damage property |  |  |  |  |
| Not willing | 5911 | 85.4 (84.4, 86.4) | 68.6 (67.6, 69.6) | 177.1 (174.6, 179.7) |
| Somewhat willing | 599 | 9.8 (9, 10.7) | 6.9 (6.4, 7.5) | 18 (16.6, 19.3) |
| Very willing | 127 | 2.5 (2, 3) | 1.5 (1.2, 1.7) | 3.8 (3.1, 4.5) |
| Completely willing | 80 | 1.4 (1.1, 1.8) | 0.9 (0.7, 1.1) | 2.4 (1.9, 2.9) |
| Non-response | 51 | 0.9 (0.7, 1.2) | 0.6 (0.4, 0.8) | 1.5 (1.1, 1.9) |
| To threaten or intimidate a person |  |  |  |  |
| Not willing | 6016 | 86.8 (85.8, 87.8) | 69.8 (68.8, 70.8) | 180.3 (177.8, 182.8) |
| Somewhat willing | 553 | 9.4 (8.6, 10.3) | 6.4 (5.9, 6.9) | 16.6 (15.2, 17.9) |
| Very willing | 77 | 1.6 (1.2, 2) | 0.9 (0.7, 1.1) | 2.3 (1.8, 2.8) |
| Completely willing | 66 | 1.2 (0.9, 1.6) | 0.8 (0.6, 0.9) | 2 (1.5, 2.5) |
| Non-response | 56 | 1.0 (0.7, 1.3) | 0.6 (0.5, 0.8) | 1.7 (1.2, 2.1) |
| To injure a person |  |  |  |  |
| Not willing | 6110 | 88.5 (87.5, 89.4) | 70.9 (69.9, 71.8) | 183.1 (180.6, 185.6) |
| Somewhat willing | 447 | 7.7 (6.9, 8.5) | 5.2 (4.7, 5.7) | 13.4 (12.2, 14.6) |
| Very willing | 82 | 1.6 (1.3, 2.1) | 1 (0.7, 1.2) | 2.5 (1.9, 3) |
| Completely willing | 63 | 1.1 (0.9, 1.5) | 0.7 (0.6, 0.9) | 1.9 (1.4, 2.4) |
| Non-response | 66 | 1.1 (0.8, 1.4) | 0.8 (0.6, 0.9) | 2 (1.5, 2.5) |
|  |  | Among respondents asked the question (n= 6,768)* | Among the full sample (n= 8,620)** | N (95% CI) (in millions) |
| To kill a person |  |  |  |  |
| Not willing | 6300 | 91.9 (91.1, 92.7) | 73.1 (72.1, 74) | 188.8 (186.4, 191.2) |
| Somewhat willing | 253 | 4.4 (3.8, 5) | 2.9 (2.6, 3.3) | 7.6 (6.7, 8.5) |
| Very willing | 80 | 1.5 (1.1, 1.9) | 0.9 (0.7, 1.1) | 2.4 (1.9, 2.9) |
| Completely willing | 79 | 1.2 (1, 1.6) | 0.9 (0.7, 1.1) | 2.4 (1.8, 2.9) |
| Non-response | 56 | 1.1 (0.8, 1.4) | 0.6 (0.5, 0.8) | 1.7 (1.2, 2.1) |

\*Respondents (n = 6,768) who considered violence to be at least sometimes justified to achieve 1 or more specific political objectives.
\*\*Percentages in this column were used for extrapolation. A weighted 21.5% (95% CI 20.6%, 22.4%) of the study sample considered violence never justified for any specific political objective.

A second series concerned categories of people as potential targets of such violence, because of who those people were (Table 7). When asked, again in a situation where they thought political violence was justified, “how willing would <u>you personally</u> be to use force or violence against a person because they are…,” 8.6% of respondents were at least somewhat willing to commit violence against “an elected federal or state government official,” 7.7% against “an elected local government official,” 5.6% against “an election worker, such as a poll worker or vote counter,” 6.4% against “a public health official,” 8.7% against “a member of the military or National Guard,” 8.7% against “a police officer,” 5.8% against “a person who does not share your race or ethnicity,” 5.2% against “a person who does not share your religion,” and 6.5% against “a person who does not share your political beliefs.”

**Table 7.**
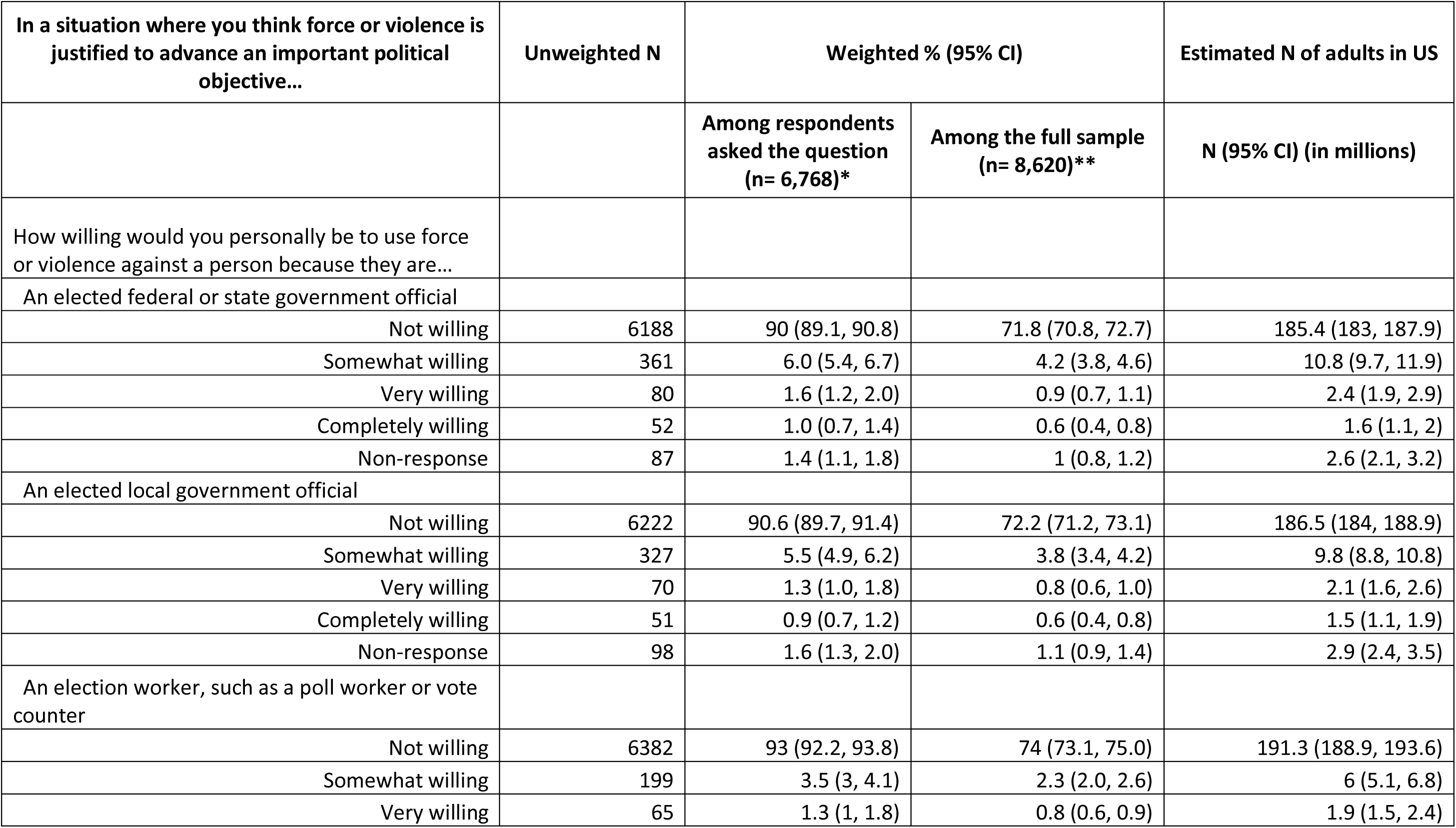
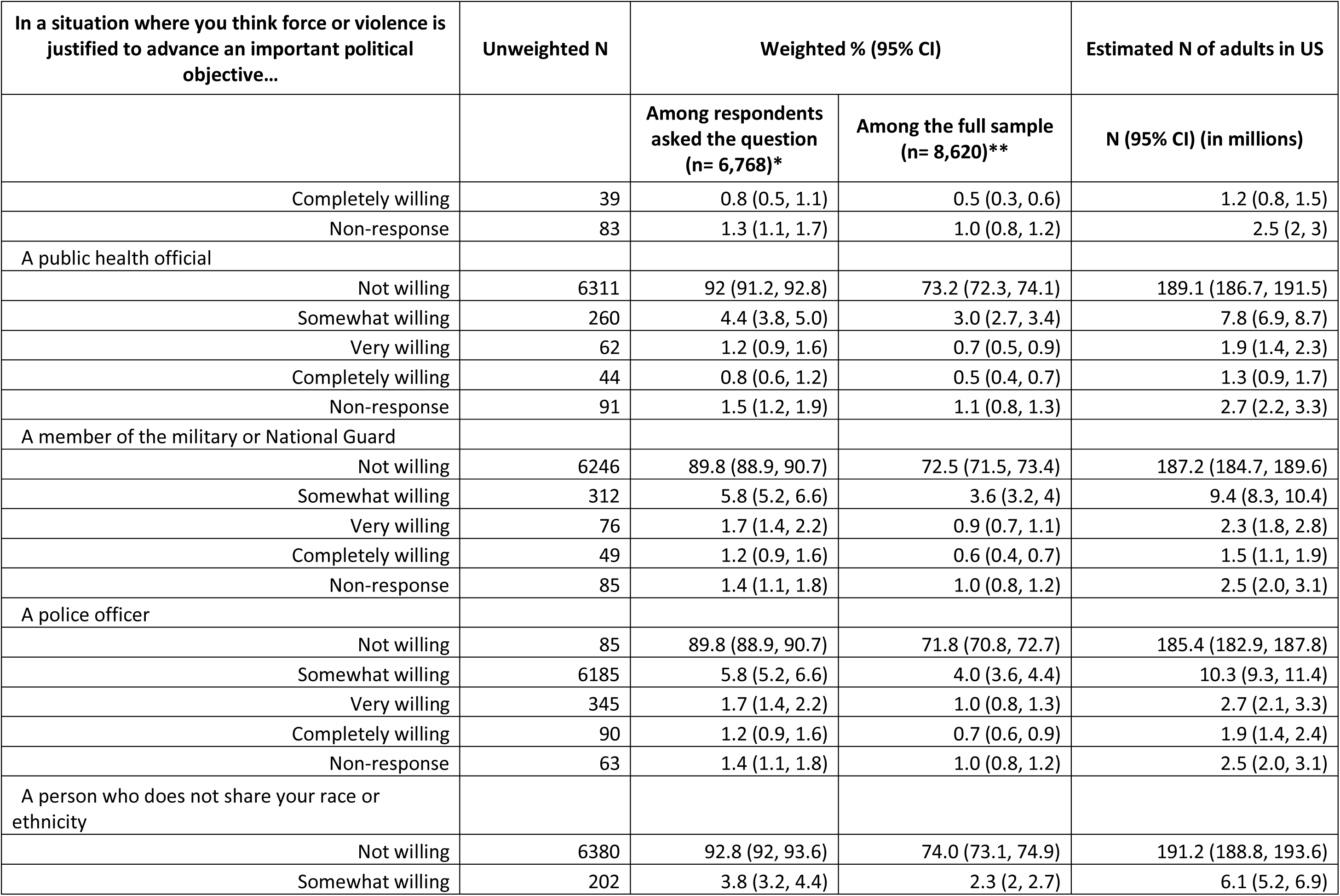
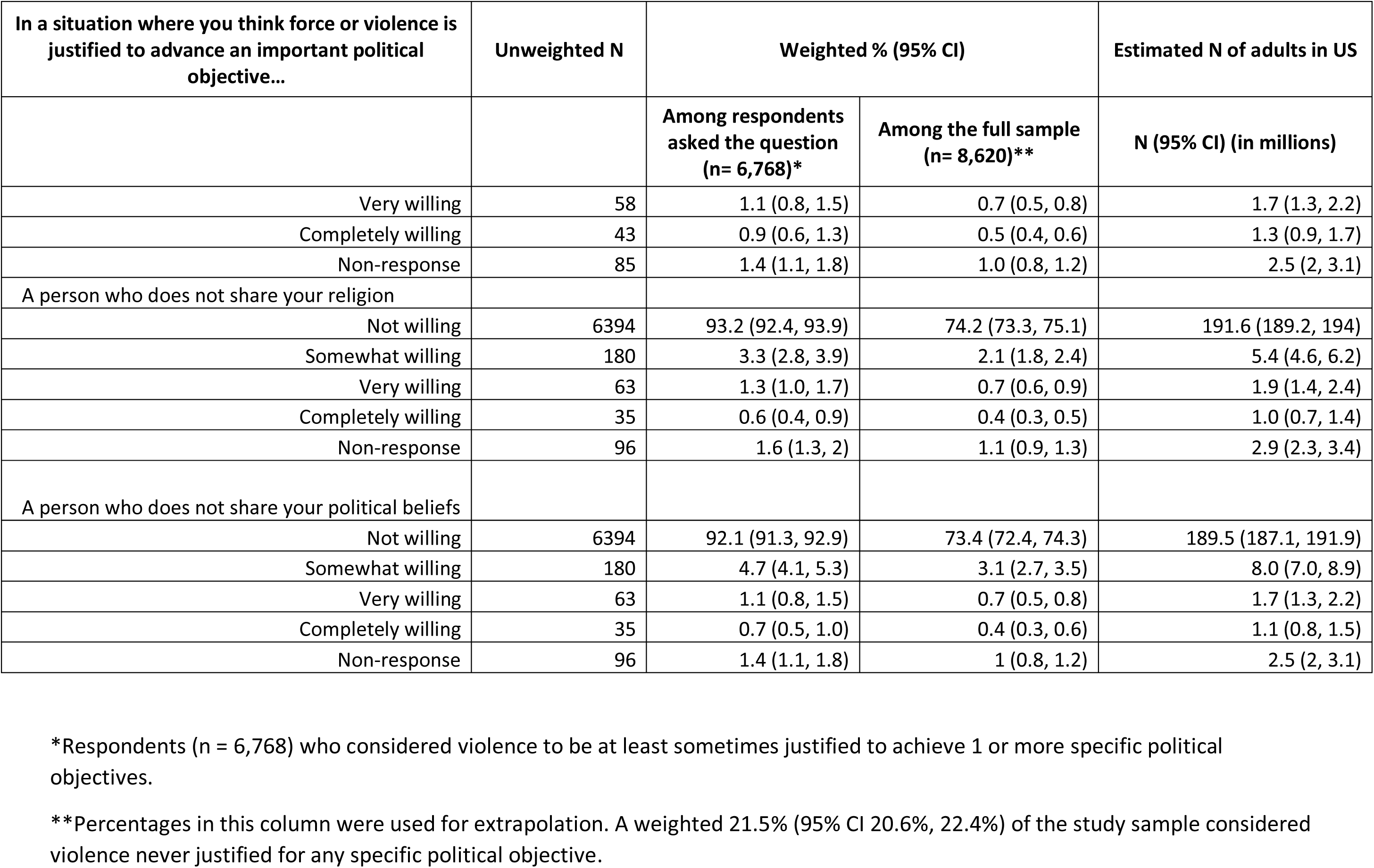
Personal willingness to engage in political violence against specific types of people among 6,768 respondents who considered violence to be at least sometimes justified to achieve 1 or more specific political objectives

| In a situation where you think force or violence is justified to advance an important political objective... | Unweighted N | Weighted % (95% CI) |  | Estimated N of adults in US |
| --- | --- | --- | --- | --- |
|  |  | Among respondents asked the question (n= 6,768)* | Among the full sample (n= 8,620)** | N (95% CI) (in millions) |
| How willing would you personally be to use force or violence against a person because they are... |  |  |  |  |
| An elected federal or state government official |  |  |  |  |
| Not willing | 6188 | 90 (89.1, 90.8) | 71.8 (70.8, 72.7) | 185.4 (183, 187.9) |
| Somewhat willing | 361 | 6.0 (5.4, 6.7) | 4.2 (3.8, 4.6) | 10.8 (9.7, 11.9) |
| Very willing | 80 | 1.6 (1.2, 2.0) | 0.9 (0.7, 1.1) | 2.4 (1.9, 2.9) |
| Completely willing | 52 | 1.0 (0.7, 1.4) | 0.6 (0.4, 0.8) | 1.6 (1.1, 2) |
| Non-response | 87 | 1.4 (1.1, 1.8) | 1 (0.8, 1.2) | 2.6 (2.1, 3.2) |
| An elected local government official |  |  |  |  |
| Not willing | 6222 | 90.6 (89.7, 91.4) | 72.2 (71.2, 73.1) | 186.5 (184, 188.9) |
| Somewhat willing | 327 | 5.5 (4.9, 6.2) | 3.8 (3.4, 4.2) | 9.8 (8.8, 10.8) |
| Very willing | 70 | 1.3 (1.0, 1.8) | 0.8 (0.6, 1.0) | 2.1 (1.6, 2.6) |
| Completely willing | 51 | 0.9 (0.7, 1.2) | 0.6 (0.4, 0.8) | 1.5 (1.1, 1.9) |
| Non-response | 98 | 1.6 (1.3, 2.0) | 1.1 (0.9, 1.4) | 2.9 (2.4, 3.5) |
| An election worker, such as a poll worker or vote counter |  |  |  |  |
| Not willing | 6382 | 93 (92.2, 93.8) | 74 (73.1, 75.0) | 191.3 (188.9, 193.6) |
| Somewhat willing | 199 | 3.5 (3, 4.1) | 2.3 (2.0, 2.6) | 6 (5.1, 6.8) |
| Very willing | 65 | 1.3 (1, 1.8) | 0.8 (0.6, 0.9) | 1.9 (1.5, 2.4) |
|  |  | Among respondents asked the question (n= 6,768)* | Among the full sample (n= 8,620)** | N (95% CI) (in millions) |
| Completely willing | 39 | 0.8 (0.5, 1.1) | 0.5 (0.3, 0.6) | 1.2 (0.8, 1.5) |
| Non-response | 83 | 1.3 (1.1, 1.7) | 1.0 (0.8, 1.2) | 2.5 (2, 3) |
| A public health official |  |  |  |  |
| Not willing | 6311 | 92 (91.2, 92.8) | 73.2 (72.3, 74.1) | 189.1 (186.7, 191.5) |
| Somewhat willing | 260 | 4.4 (3.8, 5.0) | 3.0 (2.7, 3.4) | 7.8 (6.9, 8.7) |
| Very willing | 62 | 1.2 (0.9, 1.6) | 0.7 (0.5, 0.9) | 1.9 (1.4, 2.3) |
| Completely willing | 44 | 0.8 (0.6, 1.2) | 0.5 (0.4, 0.7) | 1.3 (0.9, 1.7) |
| Non-response | 91 | 1.5 (1.2, 1.9) | 1.1 (0.8, 1.3) | 2.7 (2.2, 3.3) |
| A member of the military or National Guard |  |  |  |  |
| Not willing | 6246 | 89.8 (88.9, 90.7) | 72.5 (71.5, 73.4) | 187.2 (184.7, 189.6) |
| Somewhat willing | 312 | 5.8 (5.2, 6.6) | 3.6 (3.2, 4) | 9.4 (8.3, 10.4) |
| Very willing | 76 | 1.7 (1.4, 2.2) | 0.9 (0.7, 1.1) | 2.3 (1.8, 2.8) |
| Completely willing | 49 | 1.2 (0.9, 1.6) | 0.6 (0.4, 0.7) | 1.5 (1.1, 1.9) |
| Non-response | 85 | 1.4 (1.1, 1.8) | 1.0 (0.8, 1.2) | 2.5 (2.0, 3.1) |
| A police officer |  |  |  |  |
| Not willing | 85 | 89.8 (88.9, 90.7) | 71.8 (70.8, 72.7) | 185.4 (182.9, 187.8) |
| Somewhat willing | 6185 | 5.8 (5.2, 6.6) | 4.0 (3.6, 4.4) | 10.3 (9.3, 11.4) |
| Very willing | 345 | 1.7 (1.4, 2.2) | 1.0 (0.8, 1.3) | 2.7 (2.1, 3.3) |
| Completely willing | 90 | 1.2 (0.9, 1.6) | 0.7 (0.6, 0.9) | 1.9 (1.4, 2.4) |
| Non-response | 63 | 1.4 (1.1, 1.8) | 1.0 (0.8, 1.2) | 2.5 (2.0, 3.1) |
| A person who does not share your race or ethnicity |  |  |  |  |
| Not willing | 6380 | 92.8 (92, 93.6) | 74.0 (73.1, 74.9) | 191.2 (188.8, 193.6) |
| Somewhat willing | 202 | 3.8 (3.2, 4.4) | 2.3 (2, 2.7) | 6.1 (5.2, 6.9) |
|  |  | Among respondents asked the question (n= 6,768)* | Among the full sample (n= 8,620)** | N (95% CI) (in millions) |
| Very willing | 58 | 1.1 (0.8, 1.5) | 0.7 (0.5, 0.8) | 1.7 (1.3, 2.2) |
| Completely willing | 43 | 0.9 (0.6, 1.3) | 0.5 (0.4, 0.6) | 1.3 (0.9, 1.7) |
| Non-response | 85 | 1.4 (1.1, 1.8) | 1.0 (0.8, 1.2) | 2.5 (2, 3.1) |
| A person who does not share your religion |  |  |  |  |
| Not willing | 6394 | 93.2 (92.4, 93.9) | 74.2 (73.3, 75.1) | 191.6 (189.2, 194) |
| Somewhat willing | 180 | 3.3 (2.8, 3.9) | 2.1 (1.8, 2.4) | 5.4 (4.6, 6.2) |
| Very willing | 63 | 1.3 (1.0, 1.7) | 0.7 (0.6, 0.9) | 1.9 (1.4, 2.4) |
| Completely willing | 35 | 0.6 (0.4, 0.9) | 0.4 (0.3, 0.5) | 1.0 (0.7, 1.4) |
| Non-response | 96 | 1.6 (1.3, 2) | 1.1 (0.9, 1.3) | 2.9 (2.3, 3.4) |
| A person who does not share your political beliefs |  |  |  |  |
| Not willing | 6394 | 92.1 (91.3, 92.9) | 73.4 (72.4, 74.3) | 189.5 (187.1, 191.9) |
| Somewhat willing | 180 | 4.7 (4.1, 5.3) | 3.1 (2.7, 3.5) | 8.0 (7.0, 8.9) |
| Very willing | 63 | 1.1 (0.8, 1.5) | 0.7 (0.5, 0.8) | 1.7 (1.3, 2.2) |
| Completely willing | 35 | 0.7 (0.5, 1.0) | 0.4 (0.3, 0.6) | 1.1 (0.8, 1.5) |
| Non-response | 96 | 1.4 (1.1, 1.8) | 1 (0.8, 1.2) | 2.5 (2, 3.1) |

\*Respondents (n = 6,768) who considered violence to be at least sometimes justified to achieve 1 or more specific political objectives.
\*\*Percentages in this column were used for extrapolation. A weighted 21.5% (95% CI 20.6%, 22.4%) of the study sample considered violence never justified for any specific political objective.

Finally, all respondents were asked to predict the likelihood of their future use of a firearm “in a situation where you think force or violence is justified to advance an important political objective” (Table 8). Nearly 1 in 5 (18.5%) thought it at least somewhat likely that “I will be armed with a gun,” 9.9% that “I will carry a gun openly, so that people know I am armed,” 2.4% that “I will threaten someone with a gun,” and 4.0% that “I will shoot someone with a gun.”

**Table 8.** Anticipated use of a firearm in situations where political violence is perceived as justified

| Thinking now about the future and all the changes it might bring, how likely is it that you will use a gun in any of the following ways in the next few years—in a situation where you think force or violence is justified to advance an important political objective? | Respondents (n= 8,620) |  | Estimated N of adults in US |
| --- | --- | --- | --- |
|  | Unweighted N | Weighted % (95% CI) | N (95% CI) (in millions) |
| I will be armed with a gun. |  |  |  |
| Not likely | 7,107 | 80.1 (79.1, 81.1) | 206.9 (204.3, 209.5) |
| Somewhat likely | 833 | 10.8 (10.1, 11.6) | 27.9 (26.0, 30.0) |
| Very likely | 254 | 3.4 (3.0, 3.9) | 8.8 (7.7, 10.1) |
| Extremely likely | 318 | 4.3 (3.8, 4.8) | 11.0 (9.8, 12.5) |
| Non-response | 108 | 1.4 (1.1, 1.7) | 3.6 (2.8, 4.4) |
| I will carry a gun openly, so that people know I am armed. |  |  |  |
| Not likely | 7,779 | 88.7 (87.8, 89.5) | 229.0 (226.8, 231.1) |
| Somewhat likely | 435 | 5.7 (5.1, 6.3) | 14.7 (13.2, 16.3) |
| Very likely | 163 | 2.2 (1.8, 2.6) | 5.6 (4.7, 6.6) |
| Extremely likely | 126 | 2.0 (1.6, 2.4) | 5.1 (4.2, 6.2) |
| Non-response | 117 | 1.5 (1.2, 1.9) | 3.9 (3.1, 4.9) |
| I will threaten someone with a gun. |  |  |  |
| Not likely | 8,351 | 96.2 (95.6, 96.6) | 248.4 (247.0, 249.6) |
| Somewhat likely | 93 | 1.4 (1.1, 1.7) | 3.5 (2.8, 4.4) |
| Very likely | 38 | 0.7 (0.5, 0.9) | 1.7 (1.2, 2.4) |
| Extremely likely | 23 | 0.3 (0.2, 0.5) | 0.8 (0.5, 1.3) |
| Non-response | 115 | 1.5 (1.2, 1.9) | 3.9 (3.1, 4.9) |
| I will shoot someone with a gun. |  |  |  |
| Not likely | 8,235 | 94.6 (94.0, 95.2) | 244.4 (242.8, 245.9) |
|  | Unweighted N | Weighted % (95% CI) | N (95% CI) (in millions) |
| Somewhat likely | 198 | 2.8 (2.4, 3.3) | 7.2 (6.2, 8.5) |
| Very likely | 36 | 0.6 (0.4, 0.8) | 1.4 (1.0, 2.1) |
| Extremely likely | 40 | 0.6 (0.4, 0.8) | 1.4 (1.0, 2.0) |
| Non-response | 111 | 1.5 (1.2, 1.8) | 3.9 (3.1, 4.6) |

## DISCUSSION

The motivating premises for this survey were that current conditions in the US create both perceived threats and actual threats to its future as a free and democratic society. The findings bear out both premises. As to the former, more than two-thirds of respondents perceived “a serious threat to our democracy”; just over half expect civil war in the next few years. As to the latter, 10% thought it at most somewhat important for the US to remain a democracy; more than 40% agreed that “having a strong leader for America is more important than having a democracy”; and 20.5% considered political violence to be in general at least sometimes justified.

Many findings from this survey are concordant with those of polls taken over the last 2 years.^5–10, 26–30^ These include support by substantial minorities of the population for broad statements of the potential need for violence to save a society that has somehow headed in the wrong direction and for false beliefs, such as the Q-Anon complex, “great replacement” thinking, and the myth that Donald Trump won the 2020 Presidential election. This concordance reflects the stability of those earlier findings and provides a foundation for the new results presented here.

Our population-level extrapolations suggest that more than 50 million adults in the US consider violence to be at least sometimes justified in general to achieve political objectives that they support. More than 60 million could at least sometimes justify violence “to preserve an American way of life based on Western European traditions”; nearly 20 million could justify violence to stop people who do not share their beliefs from voting.

These are not abstract beliefs, made without commitment. Our extrapolations suggest that to achieve a political objective that they support, 6 million Americans would be very or completely willing to damage property and between 4 million and 5 million to threaten or intimidate someone, injure them, or kill them. Between 3 million and 5 million Americans would be very or completely willing to commit violence against others because they are representatives of social institutions: government officials, election officials, health officials, members of the military or police. Three million would commit politically-motivated violence against others because of differences in race/ethnicity or religion.

For many, situations in which they consider political violence to be justified call for the use of firearms. Based on our extrapolations, nearly 20 million Americans think it very or extremely likely that they will be armed in such a situation in the next few years, nearly 11 million that they will carry a gun openly, and nearly 3 million that they will shoot someone.

In the aggregate, these initial findings suggest a continuing alienation from and mistrust of American democratic society and its institutions, founded in part on false beliefs. They suggest a high level of support for violence, including lethal violence, to achieve political objectives. The prospect of large-scale political violence in the near future is entirely plausible.^31^ Forthcoming analyses will shed light on factors associated with that support and inform efforts to prevent that prospect from being realized.

It is important to emphasize that these findings also provide firm ground for hope. A large majority of respondents rejected political violence altogether, whether generally or in support of any single specific objective. A large majority of those who did endorse political violence were unwilling to resort to violence themselves. The challenge now for those large majorities is to recognize the threat posed by those who are willing to engage in political violence and respond adequately to it.

### Limitations

Several technical limitations exist. The findings are cross-sectional and subject to sampling error and bias due to nonresponse and other factors. Many important outcomes are uncommon, with response counts <100 and weighted prevalences below 5%. The large study sample results in relatively narrow confidence intervals in these cases, but the estimates remain particularly vulnerable to bias from sources such as inattentive or strategic responses. Widely publicized mass shootings occurred in Buffalo, NY and Uvalde, TX while the survey was in the field. The Buffalo shooting is understood to have been a race-related hate crime motivated by “replacement” thinking and may have affected respondents’ views on race, violence, and that particular belief. Russia’s war against Ukraine may have influenced responses on violence and democracy.

This initial report presents only simple descriptive tabulations to establish prevalences. Further analyses of these data are in progress to explore variation in those prevalences across the study population, such as with demographics, position on the political spectrum, and firearm ownership. Follow-up studies are in development to explore the meaning and implications of the findings here. For example, no questions were asked to obtain respondents’ opinions in cases where they would be helpful: does a respondent who expects civil war view that war positively or negatively? Similarly, this survey did not solicit specific information on what gives rise to support for political violence, or on how that support or its causes might best be addressed in prevention efforts.

## Conclusion

Findings from this large, nationally representative survey suggest that current conditions in the United States put at risk the future of the country as a free and democratic society. Among these are support by substantial minorities of the population for violence, including lethal violence, to obtain political objectives. Efforts to prevent that violence should proceed rapidly based on the best evidence available, while further research identifies factors associated with support for political violence and informs further prevention efforts.

## Data Availability

Data produced in the present study are subject to additional analyses and will be made available upon reasonable request to the authors when analyses are completed.

## ACKNOWLEDGMENTS

The authors gratefully acknowledge the contributions of Amanda Aubel, MPH; Deborah Azrael, PhD; Angela Bayer, PhD, MHS; Pamela Keach, MS; Nicole Kravitz-Wirtz, PhD, MPH; Matthew Miller, MD, ScD; and Rocco Pallin, MPH.

