## Supplemental text, table, and figures for "Views of American Democracy and Society and Support for Political Violence: First Report from a Nationwide Population-Representative Survey"

This manuscript is a preprint; it has not undergone peer review. Submitted to medRxiv July 15, 2022.



| Page | Title |
|---|---|
| 2 | Questions from the 2022 American Life Survey that supplied data for this study |
| 8 | References for the question list |
| 9 | Table S1. Comparison of respondents and nonrespondents |
| 11 | Figure S1. Observed and expected monthly counts of National Instant Criminal Background Check System background checks for firearm purchases, January 2014-June 2022 |
| 12 | Figure S2. Justifiability of use or force or violence to achieve specific political objectives |

**Questions from the 2022 American Life Survey that supplied data for this study**

For questions that presented a series of items for separate consideration, such as the third question below, all those items are listed, even though not all were used in this analysis.

Response options are presented here in order from negative to positive (e.g., “not important” to “extremely important”). Respondents were randomized 1:1 to receive responses in that order or the reverse.

In the list below, questions or items that were repeated or adapted from prior surveys contain citations to those surveys.

**Domain 1: democracy in the United States**

*Now we’d like to ask you a few questions about the United States as you see it now, in 2022.*

**Q:** When thinking about democracy in the United States these days, do you believe…?[1]

1. There is a serious threat to our democracy.
2. There may be a threat to our democracy, but it is not serious.
3. There is no threat to our democracy.

**Q:** How important do you think it is for the United States to remain a democracy?[2]

1. Not important
2. Somewhat important
3. Very important
4. Extremely important

**Q:** How much do you agree or disagree with the following statements about democracy in the United States?

a. Democracy is the best form of government.[3]

b. These days, American democracy only serves the interests of the wealthy and powerful.[4]

c. Having a strong leader for America is more important than having a democracy.

d. If elected leaders will not protect American democracy, the people must do it themselves, even if it requires taking violent actions.[4]

e. In the next few years, there will be civil war in the United States.[5]

1. Do not agree
2. Somewhat agree
3. Strongly agree
4. Very strongly agree

**Domain 2: American society and institutions**

*The next few questions are about your views of American society.*

**Q:** How much do you agree or disagree with each of the following statements about people in America today?

a. White people benefit from advantages in society that Black people do not have.[4]

b. Discrimination against whites is as big a problem as discrimination against Blacks and other minorities.[4]

c. Our American way of life is disappearing so fast that we may have to use force to save it.[4]

d. In America, native-born white people are being replaced by immigrants.

e. Having more Black Americans, Latinos, and Asian Americans is good for the country.[6]

1. Do not agree
2. Somewhat agree
3. Strongly agree
4. Very strongly agree

**Q:** People have many different views about American society. How much do you agree or disagree with each of the following?

a. The government, media, and financial worlds in the U.S. are controlled by a group of Satan-worshipping pedophiles who run a global child sex trafficking operation.[7]

b. There is a storm coming soon that will sweep away the elites in power and restore the rightful leaders.[7]

c. Because things have gotten so far off track, true American patriots may have to resort to violence in order to save our country.7

d. The chaos in America today is evidence that we are living in what the Bible calls "the end times."[8]

e. Capitalism is a system of oppression and should be abolished.

f. Straight white men hold far too much power in America.

g. The 2020 election was stolen from Donald Trump, and Joe Biden is an illegitimate president.

h. Armed citizens should patrol polling places at election time.

1. Do not agree
2. Somewhat agree
3. Strongly agree
4. Very strongly agree

**Domain 3: violence, including political violence**

*Now we have a few questions about the use of force or violence. A reminder: your responses will be kept confidential and anonymous*.

**Q:** In general, what do you think about the use of force or violence in the following situations—is it never justified, sometimes justified, usually justified, or always justified? "Force or violence" means physical force strong enough that it could cause pain or injury to a person.

(Not randomized)

a. In self defense

b. To prevent someone from injuring or killing another person

c. To prevent someone from injuring or killing themselves

d. To prevent harm or damage to property

e. To win an argument

f. In response to an insult

g. To get respect

1. Never justified
2. Sometimes justified

3. Usually justified
4. Always justified

**Q:** People sometimes talk about using force or violence to achieve political objectives. In general, what do you think about using force or violence to advance an important political objective that you support—is it…?

1. Never justified
2. Sometimes justified
3. Usually justified
4. Always justified

**Q:** Again, your view of the use of force or violence to advance an important political objective might depend on the specific objective that was involved. What do you think about the use of force or violence in the following situations—is it never justified, sometimes justified, usually justified, or always justified?

a. To return Donald Trump to the presidency this year

b. To stop an election from being stolen

c. To stop people who do not share my beliefs from voting

d. To prevent discrimination based on race or ethnicity

e. To preserve an American way of life based on Western European traditions

f. To oppose the government when it does not share my beliefs

g. To oppose the government when it tries to take private land for public purposes

1. Never justified
2. Sometimes justified
3. Usually justified
4. Always justified

*The next questions are about your personal willingness to use force or violence.*

**(Questions asked of respondents who endorsed at least 1 use of violence to achieve a specific political objective.)**

**Q:** In a situation where you think force or violence is justified to advance an important political objective, how willing would you personally be to use force or violence in each of these ways?

a. To damage property

b. To threaten or intimidate a person

c. To injure a person

d. To kill a person

1. Not willing
2. Somewhat willing
3. Very willing
4. Completely willing

**Q:** In a situation where you think force or violence is justified to advance an important political objective, how willing would you personally be to use force or violence against a person because they are…

a. An elected federal or state government official

b. An elected local government official

c. A public health official

d. A member of the military or National Guard

e. A police officer

f. A person who does not share your race or ethnicity

g. A person who does not share your religion

h. An election worker, such as a poll worker or vote counter

i. A person who does not share your political beliefs

1. Not willing

2. Somewhat willing
3. Very willing
4. Completely willing

**(Question asked of all respondents.)**

**Q:** Thinking now about the future and all the changes it might bring, how likely is it that you will use a gun in any of the following ways in the next few years—in a situation where you think force or violence is justified to advance an important political objective?

a. I will be armed with a gun.

b. I will carry a gun openly, so that people know I am armed.

c. I will threaten someone with a gun.

d. I will shoot someone with a gun.

1. Not likely
2. Somewhat likely
3. Very likely
4. Extremely likely

Table S1. Comparison of respondents and nonrespondents

Weights were not available for non-respondents; this table presents unweighted counts and percentages.

| Characteristic | Respondents (n= 8, 620) | | Nonrespondents (n= 6,099) | |
|---|---|---|---|---|
| | Unweighted N | Unweighted % | Unweighted N | Unweighted % |
| **Age** (mean [SD]) | 53.8 (17.2) | | 42.9 (16.2) | |
| 18-24 | 447 | 5.2 | 977 | 16.0 |
| 25-34 | 1024 | 11.9 | 1129 | 18.5 |
| 35-44 | 1374 | 15.9 | 1340 | 22.0 |
| 45-54 | 1215 | 14.1 | 1151 | 18.9 |
| 55-64 | 1833 | 21.3 | 811 | 13.3 |
| 65-74 | 1788 | 20.7 | 502 | 8.2 |
| 75+ | 939 | 10.9 | 189 | 3.1 |
| Non-response | 0 | 0.0 | 0 | 0.0 |
| **Gender** | | | | |
| Female | 4373 | 50.7 | 3313 | 54.3 |
| Male | 4247 | 49.3 | 2786 | 45.7 |
| Non-response | 0 | 0.0 | 0 | 0.0 |
| **Race/Ethnicity** | | | | |
| White, non-Hispanic | 6047 | 70.2 | 3528 | 57.9 |
| Black, non-Hispanic | 836 | 9.7 | 836 | 13.7 |
| Hispanic, any race | 1084 | 12.6 | 1183 | 19.4 |
| Other, non-Hispanic | 392 | 4.6 | 304 | 5.0 |
| 2+ Races, non-Hispanic | 261 | 3.0 | 248 | 4.1 |
| Non-response | 0 | 0.0 | 0 | 0.0 |
| **Marital status** | | | | |
| Now married | 5246 | 60.9 | 3128 | 51.3 |
| Widowed | 443 | 5.1 | 168 | 2.8 |
| Divorced | 909 | 10.6 | 577 | 9.5 |
| Separated | 139 | 1.6 | 141 | 2.3 |
| Never married | 1883 | 21.8 | 2085 | 34.2 |
| Non-response | 0 | 0.0 | 0 | 0.0 |
| **Education** | | | | |
| No high school diploma or GED | 542 | 6.3 | 625 | 10.3 |
| High school graduate (diploma or GED) | 2158 | 25.0 | 1759 | 28.8 |
| Some college or Associate's degree | 2364 | 27.4 | 1769 | 29.0 |
| Bachelor's degree | 1951 | 22.6 | 1140 | 18.7 |
| Master’s degree or higher | 1605 | 18.6 | 806 | 13.2 |

| Characteristic | Respondents (n= 8, 620) | | Nonrespondents (n= 6,099) | |
|---|---|---|---|---|
| | Unweighted N | Unweighted % | Unweighted N | Unweighted % |
| Non-response | 0 | 0.0 | 0 | 0.0 |
| **Household Income** | | | | |
| Less than $10,000 | 272 | 3.2 | 312 | 5.1 |
| $10,000 to $24,999 | 745 | 8.6 | 609 | 10.0 |
| $25,000 to $49,999 | 1469 | 17.0 | 1158 | 19.0 |
| $50,000 to $74,999 | 1414 | 16.4 | 1012 | 16.6 |
| $75,000 to $99,999 | 1214 | 14.1 | 810 | 13.3 |
| $100,000 to $149,999 | 1500 | 17.4 | 1031 | 16.9 |
| $150,000 or more | 2006 | 23.3 | 1167 | 19.1 |
| Non-response | 0 | 0.0 | 0 | 0.0 |
| **Employment** | | | | |
| Working full time | 3888 | 45.1 | 3377 | 55.4 |
| Working part time | 1132 | 13.1 | 1055 | 17.3 |
| Not working | 3600 | 41.8 | 1667 | 27.3 |
| Non-response | 0 | 0.0 | 0 | 0.0 |
| **Census region** | | | | |
| New England | 412 | 4.8 | 258 | 4.2 |
| Mid-Atlantic | 1090 | 12.7 | 747 | 12.3 |
| East-North Central | 1267 | 14.7 | 840 | 13.8 |
| West-North Central | 604 | 7.0 | 420 | 6.9 |
| South Atlantic | 1714 | 19.9 | 1232 | 20.2 |
| East-South Central | 465 | 5.4 | 432 | 7.1 |
| West-South Central | 904 | 10.5 | 840 | 13.8 |
| Mountain | 745 | 8.6 | 426 | 7.0 |
| Pacific | 1419 | 16.5 | 904 | 14.8 |
| Non-response | 0 | 0.0 | 0 | 0.0 |

Figure S1. Observed and expected monthly counts of National Instant Criminal Background Check System background checks for firearm purchases, January 2014 to June 2022

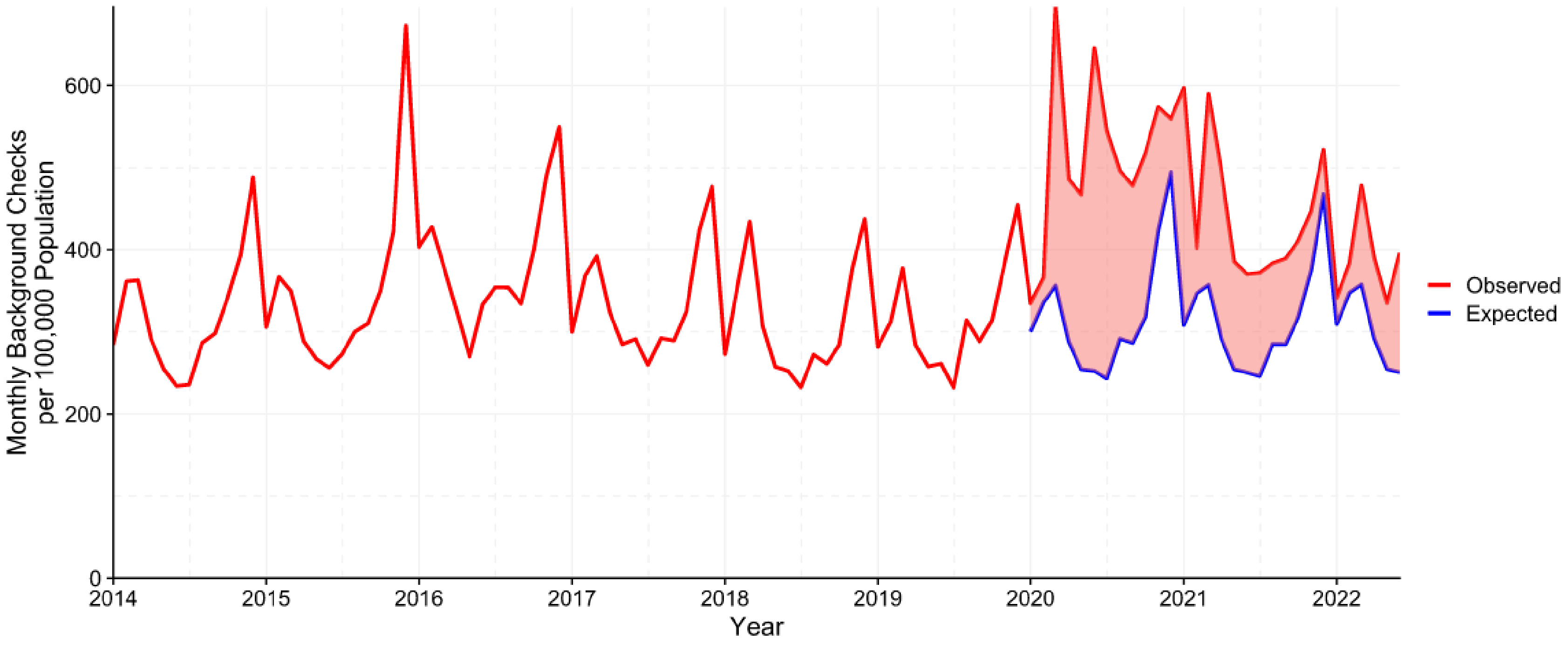


Expected counts for 2020 to 2022 were obtained by fitting an ARIMA model to data for January 2007 through December 2019.

Figure S2. Justifiability of use or force or violence to achieve specific political objectives

Respondents (n= 8,620) were asked, “What do you think about the use of force or violence in the following situations?” with response options always/usually/sometimes/never justified.

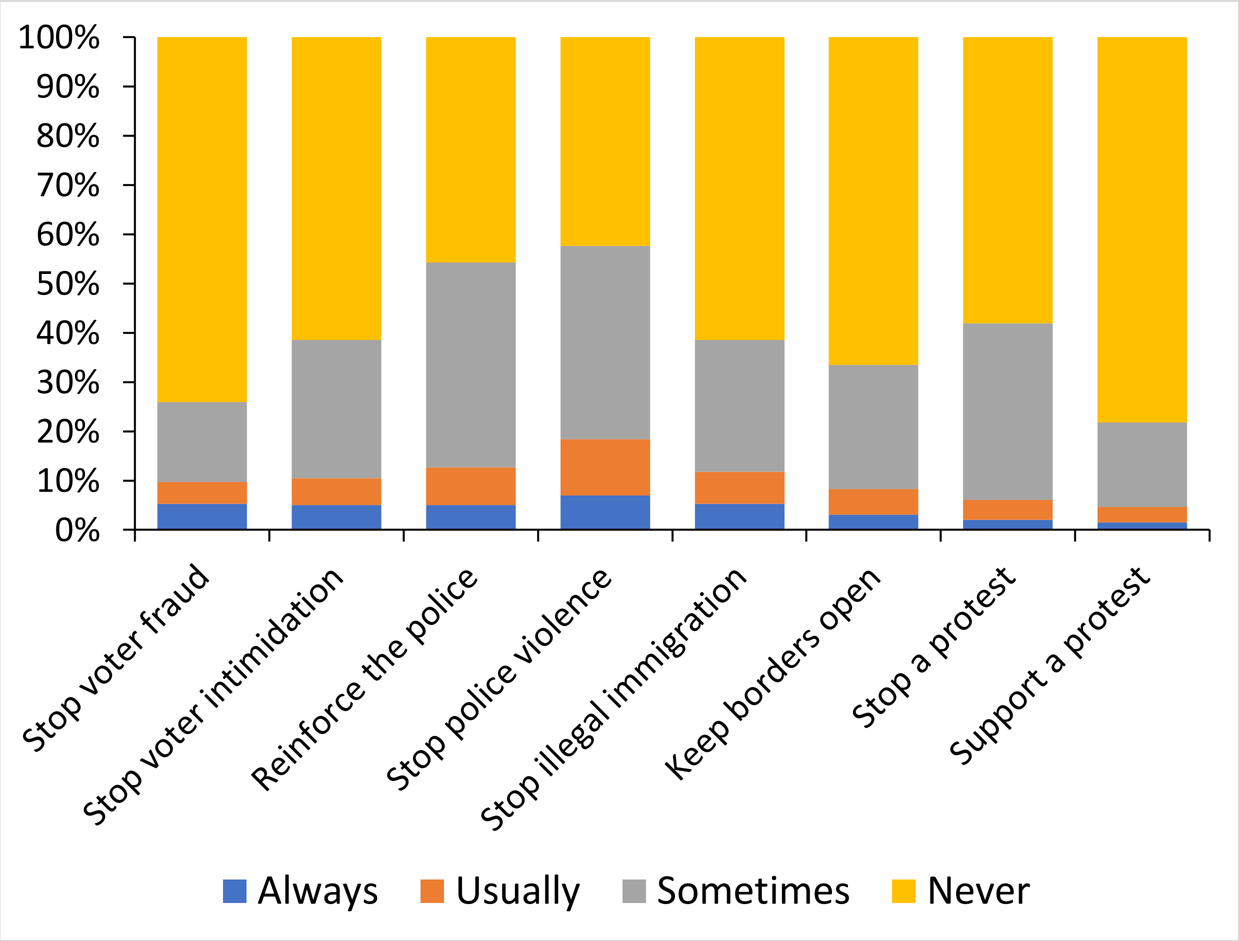
